# Integrating mental health support into care for placenta accreta spectrum: A qualitative analysis of patient perspectives

**DOI:** 10.64898/2026.07.02.26356969

**Authors:** Natalie Feldman, Margo D Nathan, Jessica M Lipschitz, Kate Salama, Lorna Campbell, Penny Wang, Leena Mittal, Daniela A. Carusi

**Author notes:** Corresponding author: Natalie Feldman.

## Abstract

**Background:** Patients with high-risk pregnancies due to placenta accreta spectrum (PAS) are at high risk of morbidity and mortality, which may increase risk for childbirth related mental health sequelae including postpartum post-traumatic stress disorder (PTSD) and trauma symptoms. However, there has been limited investigation into these patients’ mental health needs. We aimed to use qualitative data to understand PAS patients’ mental health experiences through their obstetric course, and to generate recommendations for the delivery of mental health support to these patients.

**Methods:** This exploratory study used a focus group format with patients who had a history of PAS. General questions about patient’s pregnancies, births, and postpartum experiences were asked by mental health professionals. Using a rapid qualitative analysis approach, transcriptions of these focus groups were coded by three psychiatrists and core themes were extracted.

**Results:** We conducted four focus groups with a total of 22 women. Major emotional themes included fear and isolation during the antepartum period, and grief, anxiety, and trauma in the postpartum period. Both periods were associated with a negative emotional impact on relationships with family members. Sadness & depression were less prominent among participants’ experiences. Participants felt that mental health care resources needed to be integrated with their obstetric care, extend further into the postpartum period, and should be as specific as possible to their medical condition.

**Conclusions:** Based on the results of these focus groups, we propose that patients with high-risk pregnancies and/or a history of traumatic birth should have access to expert mental health care that is integrated with their obstetric care. These patients may benefit from extended obstetric follow-up. Mental health screening in these populations should focus on anxiety and trauma symptoms rather than only screening for depression. Future studies should continue to examine these factors in a broader group of women with high-risk pregnancies beyond PAS.

## Background

Nearly half of women perceive their childbirth as traumatic, making them more vulnerable to the development of perinatal psychiatric illnesses, including anxiety, depression, and post-traumatic stress disorder.^1,2^ Further, more medically complex pregnancies have been independently identified as a risk factor for the development of perinatal psychiatric illness.^3,4^ Postpartum post-traumatic stress disorder (PTSD) has been identified in 4% of birthing patients and 19% of high-risk obstetric patients, emphasizing the impact of trauma in complicated pregnancies.^1,5^ In addition to general pregnancy complications, risk factors for a traumatic birth experience include operative and emergency births, postpartum hemorrhage, post-surgical pain, negative perceptions of the birth experience, and lack of labor support.^2,6,7^ Low social support, dissatisfaction with healthcare experiences, and fear of future childbirth proximal to delivery predict a higher burden of post-traumatic symptoms^8^, providing insight into potential points for intervention to decrease risk. Interventions may include early identification and support of those at high risk for negative psychological impact, calm and consistent communication during pregnancy and delivery, early skin-to-skin contact between mother and baby, and extended follow-up over the first year postpartum with the opportunity to review the events of delivery.^7,9,10^ Further, screening for perinatal depression, anxiety, and trauma related disorders as well as encouraging mental health support regardless of diagnosis have also been recommended in prior studies.^10,11^

Placenta accreta spectrum (PAS) is one of the strongest known risk factors for severe maternal morbidity and mortality at delivery, mediated through major hemorrhage and hysterectomy as well as other delivery and postpartum complications.^12^ While multiple studies have shown a link between emergency hemorrhage, peripartum hysterectomy and PTSD.^13–15^ PAS stands out in that these risks are often anticipated during pregnancy. This can theoretically extend the potential for emotional distress but may also provide opportunities for intervention and mitigation. Despite the significant morbidity and mortality associated with PAS, there has been little exploration of the psychological experiences of women with this condition or investigation into how to best support them. Limited prior research examining the experience of patients with PAS has identified themes of fear, trauma, lack of autonomy, and difficult coping. Regarding mental health support, these studies emphasized the need for patient counseling and referrals, as well as obstetric care systems designed to address patients’ emotional needs.^16–18^ The standard model for PAS care involves multidisciplinary teams, including experts in obstetrics, surgery, imaging, transfusion, critical care, anesthesiology and neonatology.^12,19^ Mental health support has been identified as an important domain, though to date there has been little investigation into the types of mental health services patients with PAS prefer and little specific guidance on creating systems for this care delivery.

In this study we aimed to qualitatively characterize both the psychological experiences of patients with PAS and the types of services that those patients want. Our exploration of desired services focused on identifying aspects of patient care that were perceived as supportive and aspects that were perceived as needing improvement. Results can inform care delivery models for these patients, as well as for others facing complicated and potentially traumatic birth experiences.

## Methods

This qualitative study was conducted in a focus group format, with Institutional Review Board approval obtained from Mass General Brigham Healthcare (IRB#2018P000307). Informed consent was obtained from participants before each focus group session started, including consent for audio recording.

### Research Team

The research team included faculty and staff from both the Obstetrics and Gynecology and Psychiatry Departments at Brigham and Women’s Hospital (BWH) with prior clinical experience working with patients with PAS. One author, JL, was a health services researcher. All focus group interviews were conducted by two authors (LM and LC) who had experience moderating group sessions. One author (DC) had provided clinical care to the patients, and thus did not participate in focus groups, have direct contact with participants, have access to identified transcripts, or engage in qualitative analysis prior to the consensus-building stage.

### Sample Selection and Study Site

Potential participants were identified by electronic medical record review of women with a clinical or pathologic diagnosis of PAS who delivered at BWH between 2008-2018. Patients were included if PAS had been suspected during the antepartum period or if there had been a major hemorrhage or hysterectomy. Eligibility criteria otherwise included English-speaking patients 18 years old and over. Patients who delivered within twelve months of study recruitment, were currently pregnant, or experienced a fetal or neonatal demise were excluded from participation. The recruitment period was from October 2018-June 2019. Potential participants were mailed a postcard describing the study and invited to contact the study team if they were interested in participating. Those who did not respond were called and offered more information. Interested patients were scheduled into planned focus groups with an intended size of 7-12 subjects.

Focus groups were conducted in-person at a hospital-affiliated center located three miles from the site of delivery. This location was chosen to mitigate possible emotional impact associated with the original location of their PAS clinical care. The focus group questions that were developed for this study can be seen in the supplement to this manuscript.

### Data Collection

One-hour focus group sessions were conducted in-person, audio-recorded, and transcribed later. At the start of each focus group session, participants completed a brief anonymous survey which covered demographics, core features of their delivery, and their personal experiences with mental health services. Interviewers LM and LC then followed a semi-structured interview guide developed and reviewed by the author group prior to the first session. The interview began with open-ended questions to generate discussion including 1) What were the aspects of care during pregnancy that were helpful and 2) Are there things that could have been added to your care that would have been helpful to your emotional wellbeing? Additional questions in the semi-structured interview guide included 3) What are the impacts that your complicated pregnancy and delivery, had on your mental and emotional wellbeing? 4)Would you have been interested in mental health care as part of your obstetric care -either during or after birth? 5) Did you find resources outside your care at BWH that were particularly helpful (such as other providers, groups, Internet resources, peer/family)? 6) In addition to the medical issues, where there were other factors that were particularly stressful during her pregnancy, or contributed to the stress of your experience? 7) How was your family impacted by the experience you had during her pregnancy and delivery? Participants received refreshments as well as $75 for their time and effort. Data were preliminarily reviewed following each session to determine if future sessions were needed or if data saturation was reached.

### Coding and Analysis

A rapid qualitative analysis approach was used^20,21^ to identify themes of participants’ emotional experience of having a high-risk pregnancy with PAS and to identify important aspects of treatment preferences. First, study team members (NF, MN, KS) created a templated summary of each focus group transcript. These summaries were organized by codes that aligned with questions from the interview guide. We also noted any emergent codes or unique concepts presented in focus group transcripts and identified representative quotations for each code. Summaries were then used to create a matrix with codes as columns and rows for each focus group conducted. Study team members (NF, MN, KS) independently reviewed this matrix to identify themes within each code based on repetition and emphasis. The full authorship team attended a series of meetings to reach consensus on included themes. This method of qualitative analysis is designed for health services research questions and has been found to yield comparable results to traditional qualitative approaches.^21,22^

## Results

### Overview

Our sample included 22 women across four focus groups. Demographics of the study participants are displayed in Table 1. Sixteen out of 22 subjects (73%) delivered in the five years prior to the study. The majority of participants experienced blood transfusion (73%), hysterectomy (77%), and/or neonatal intensive care unit admission (77%). Most had an antepartum admission (59%) and delivered prior to 38 weeks gestation (77%).

**Table 1:**
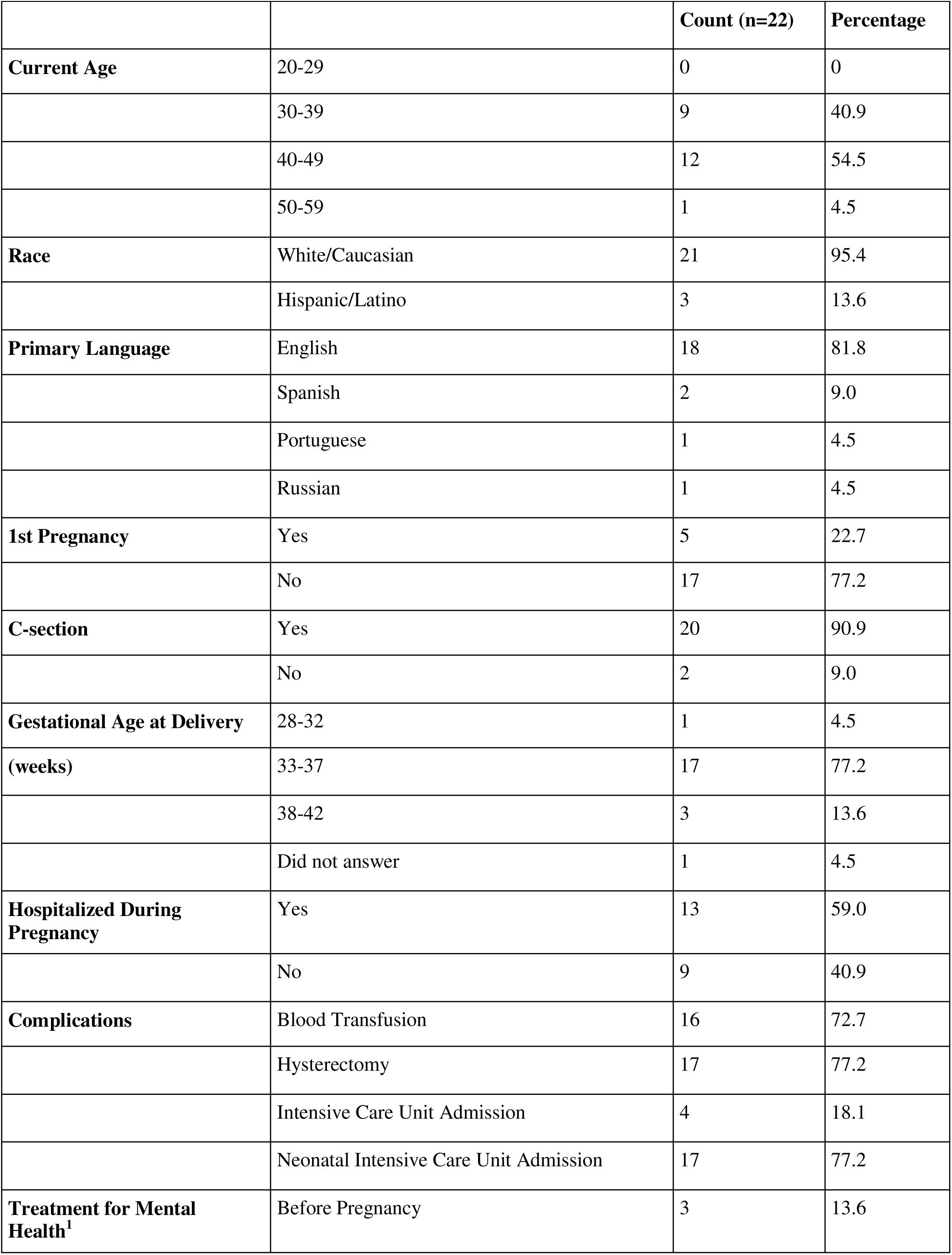
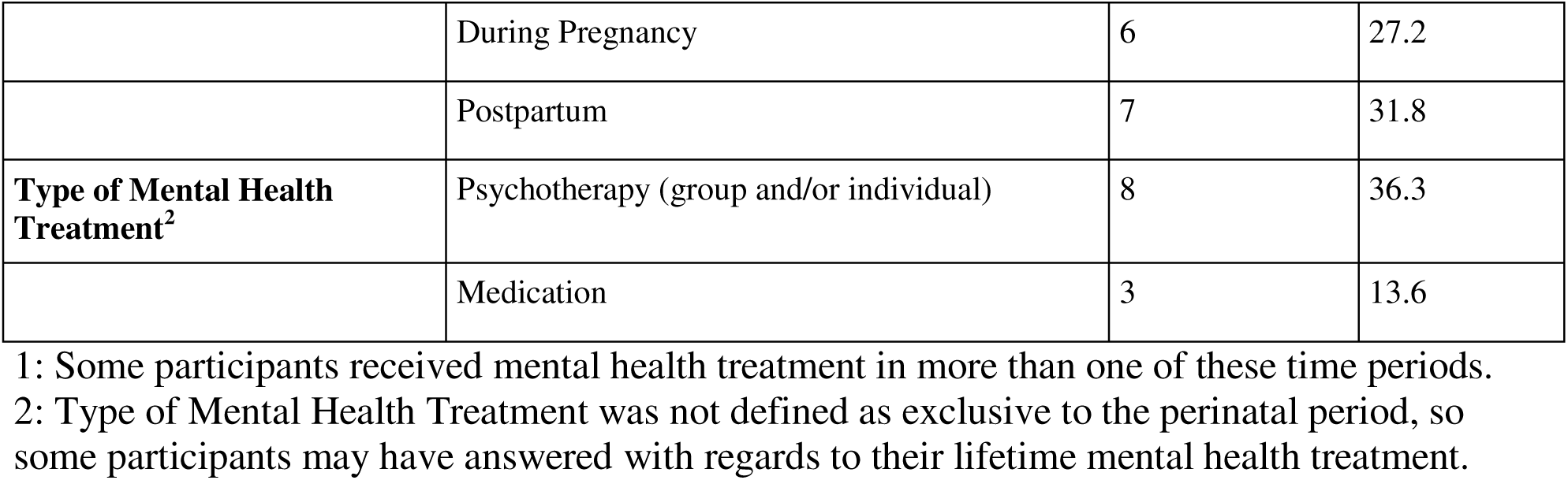
Descriptive Characteristics of Study Participants.

Key themes and subthemes are listed below, with illustrative quotations in Table 2.

**Table 2:** Quotations Illustrating Key Themes.

| **ANTEPARTUM** |  |
| --- | --- |
| *Emotional and Functional Impact - Antepartum* |  |
| *Theme* | Example Quotations |
| *Fear* | “I was scared to go out, like everywhere I went I thought ‘what if I bleed while I’m there? Where’s the nearest hospital? Do I have other clothes with me?’”<br>“I think, truthfully, if you’ve been given this diagnosis, you’re just scared.” |
| *Isolation* | “I was lonely. I mean, I lived in the hospital [during prolonged antepartum admission for PAS]. I see my husband for dinner every night. Occasionally, friends would come to visit, but basically, I was really lonely.”<br>“I think when you’re dealing with a complicated pregnancy, it feels very lonely.” |
| *Important Aspects of Care - Antepartum* |  |
| *Theme* | Example Quotations |
| *Education about diagnosis* | “I was able to sit and talk with [OB doctor] like a month before the baby was born, and she said, “Well, these are the different scenarios. This is what could happen, and I just want you to be aware, so if you are in an emergency we’re not trying to explain all the different things that could happen. “So for me, that was helpful...I just knew what to expect.”<br>“I think what could have helped is maybe really sort of talking before, even let me know that there is a chance [of needing a hysterectomy]. So make me more aware that something like that could happen.” |
| *Integrated emotional health care* | “If OB doctor or whoever had said just go to a psychiatrist, I wouldn't have gone. She said, "This is someone I work hand-in-hand with who talks to lots of accreta patients." She presented it in a very different way.” |
|  | “Really just have it be a wellbeing check, ‘it’s our policy, we’ll be checking in on you.’ Not like a ‘do you want me to’ because no, I’m not going to want you to. But if it’s like a ‘this is how our office works and we are just checking in’ and then I can choose to answer the phone or not.” |
| *Care team consistency* | “Every ultrasound technician would give me a completely different picture of what was going on with me...it was like an emotional roller coaster.”<br>“I think one of the huge supports for me were the nurses because they knew me for so long. They knew me pre-delivery. They knew me post-delivery. They were super supportive.” |
| *Provider expertise in diagnosis* | “The fact that she wasn’t just any psychiatrist, she knew about [my condition]...I didn’t have to explain it to her. It seemed like she had had similar discussions with other patients before, and I just saw her throughout my pregnancy at the end. It was super helpful.”<br>“I trust my doctor...she is the specialist. She know [sic] what she is doing.” |
| *Agency in decision-making* | “There was only one outcome for me, which was going to be a hysterectomy, but she presented it in a way that made me feel like I had a choice...just psychologically, I felt better having that sense of control...I felt like it was a decision I chose, and I was okay moving on after that.”<br>“I was still able to breastfeed because I was like that’s really a priority for me, and they were very respectful for that, and I can’t imagine all the things they had to figure out to make that happen, but they were great with that.” |
| **POSTPARTUM** |  |
| *Emotional and Functional Impact – Postpartum* |  |
| *Theme* | *Example Quotations* |
| *Grief* | “I almost wish there were grief counseling...you’re alive and your baby’s alive, but grieving whatever your life was before, as well as grieving how your body’s changed.”<br>“Now it’s more of the grieving my third child that I can’t have, and my organs.” |
| *Anxiety* | “[after delivery] that’s when a lot of the angst and anxiety from the pregnancy came out afterwards...it all fell apart.”<br>“I didn’t want to leave the house. And in some ways I couldn’t leave the house because my daughter was premature and I was scared to death and I pumped for her for 13 months, which means I never left the house.” |
| *Trauma* | “There is so much PTSD. I’m a nurse. I can’t even look at giving blood without thinking about the transfusions that I got because it was an emergency delivery...and I’m a year and a half out, but still, it’s still there.”<br>“I have vivid memories of someone rushing in with a bag of blood and it breaking all over the floor where I was watching. I was diagnosed with post-traumatic stress disorder.” |
| *Avoidance of hospital/medical care* | “I said “Oh, no, no, no. I’m never coming back here. I’m done with [Hospital where I delivered]. I’ve lived here, I’m good.”<br>“I was like ‘Well, I don’t have a uterus anymore. I don’t even really want to go back to the OB. I don’t even want to back to the Brigham. I kind of don’t want to go back.’ So the thought of me actually going in there and saying ‘I need help’, at one year, that was more than I was capable of.” |
| *Important Aspects of Care - Postpartum* |  |
| *Theme* | *Example Quotations* |
| *Minimize separation from baby* | “I think for me that was the most traumatic thing. It was like two and a half days before I got to hold my baby.”<br>“My baby ended up going to the NICU for a while because he had breathing apnea. I was really bummed about that.” |
| *Longer postpartum mental health follow-up* | “In the immediate, you’re just like ‘Oh, I’m alive. All right. This is great.’...But then it’s later down the road, exactly as everyone has said, that you’re kind of like ‘Oooh, no, I’m not. It was a lot that happened in that seven, nine months’, whatever. Something feels out of control.”<br>“It’s a survivor anniversary in addition to your child’s first birthday. And that day was really |
|  | reflective for me and really hard. I cried the entire day.” |
| *Education around body changes* | “What happens after a hysterectomy? Like yeah, am I still gonna get my period? Will this affect my hormones? The attention was more on like, the C-section and the baby than it was on that part of things.”<br>“Your body has gone through this huge trauma. You’re not the same no matter how you want to look at it, and that’s never discussed. You don’t pee the same. You don’t poop the same. Intercourse isn’t--nothing’s the same. And nobody scratches that surface.” |
| **ACROSS ANTEPARTUM AND POSTPARTUM** |  |
| *Emotional and Functional Impact – Across Antepartum and Postpartum* |  |
| *Theme* | Example Quotations |
| *Negative impact on relationships/family* | “[my husband] tries to be strong. But sometimes he needs to talk about it, but I’m not the person who wants to hear about all that blood. And I don’t know, I feel like my marriage took a hit. I do...we’re still clawing our way back, and I hope we make it.”<br>“I mean, you just can’t be four years old and see your mom crying every day for months and have a really well-settled, great, confident feeling about that.” |
| *Important Aspects of Care – Across Antepartum and Postpartum* |  |
| *Theme* | Example Quotation |
| *Peer support from others with similar experiences* | “I think it’s really nice for people to have a chance to process with people that have been through this.”<br>“I would’ve only gone to [a support group]...with women who go to bed crying every night thinking they’re going to die.” |

### Emotional and Functional Impact During Antepartum Period

#### Fear

Many participants described a profound sense of fear during the antepartum period.

Fears were focused on the potential for bad outcomes, including concerns about death in childbirth and negative consequences for the baby. Multiple participants also expressed fears about bleeding during pregnancy.

#### Isolation

Participants expressed that the antepartum period was heavily marked by feelings of loneliness. More specifically, participants described being lonely during long hospital stays where they did not have access to consistent interaction with friends and family. Participants also described feeling alone in their experience of a medically complex pregnancy, because others could not understand what they were going through.

### Important Aspects of Care During Antepartum Period

#### Education About Diagnosis

Many participants reported that receiving education about their diagnosis prior to birth was helpful. Participants appreciated learning in advance about various outcomes and procedures that could occur, rather than initially hearing these things during an emergent situation. Participants also expressed that education allowed them to understand what to expect and how to prepare themselves.

#### Integrated Emotional Healthcare

Participants conveyed that they appreciated the integration of a mental health professional into their OB/GYN team. They expressed that they found it helpful to have mental health care as a routine part of their obstetric care. Many indicated that they did or would have declined if they were offered a referral to an external mental health professional.

#### Provider Consistency

Many participants suggested that they experienced more stress during the antepartum period when they received different information from different care team members. Specifically, multiple participants referred to conflicting comments about their condition from different ultrasound technicians. They also endorsed stress when meeting with covering care team members or many different providers who shared differing opinions.

#### Provider Expertise in Diagnosis

Participants consistently reported that they liked meeting with providers (mental health providers and OB/GYNs) with experience working with women with their diagnosis. They expressed comfort in not having to explain their condition to a provider and trusting that their provider was knowledgeable about their condition.

#### Agency in Decision-Making

Participants communicated that it was important to them when the medical team gave them choices and respected their preferences. Some participants expressed that, even when a specific health outcome was inevitable, it was important that they received information and had an opportunity to voice a preference in a way that made them feel like they had some control.

### Emotional and Functional Impact During Postpartum Period

#### Grief

Participants described different experiences of grief and loss after their deliveries. Losses that were identified included the loss of future childbearing potential, the uterus, and other aspects of what their body was like pre-pregnancy.

#### Anxiety

Participants identified different experiences of anxiety as a prominent feature of their postpartum period. Some described feeling like the worry and “angst” of antepartum manifested strongly after the delivery. Also reported was an experience of retrospective ruminative guilt, with persistent questioning over whether they made the right choices about their delivery. Some participants also described disproportionate worry about the baby’s well-being.

#### Trauma

Several participants reported that their delivery experience was traumatic and was followed by experiences that they described as post-traumatic. Prominent symptoms included intrusive memories and/or flashbacks related to their delivery. They also noted specific triggers, including certain places, times, and cues that brought back unwanted memories of their delivery.

#### Avoidance of Hospital/Medical Care

Some participants were unable to return to their delivery hospital for follow-up care, even to ask for needed help. In many cases, this avoidance was described as part of a constellation of post-trauma symptoms. Additionally, some participants acknowledged had a prolonged antepartum hospitalization at their delivery hospital contributed to the avoidance.

### Important Aspects of Care During Postpartum Period

#### Minimize Separation from Baby

Participants expressed a strong dislike of being separated from the newborn due to NICU admission and/or maternal acuity. Participants recommended minimizing this gap.

#### Longer Postpartum Mental Health Follow-Up

Participants noted that the mental health impact of their deliveries often hit months or even a year postpartum. Some noted that their child’s first birthday was a particular milestone that brought up emotions from around their delivery.

#### Education Around Body Changes

Participants shared that their complicated deliveries (and in many cases hysterectomies) profoundly changed their physical and hormonal functioning. They indicated that they received insufficient education and preparation for these changes.

### Emotional and Functional Impact Across Antepartum and Postpartum Periods

#### Negative Impact on Relationships/Family

Multiple participants observed that their pregnancy and delivery experiences impacted their families as well. Some noted that partners also had a traumatic response to their delivery; in multiple cases, these experiences were described as damaging or ending a marriage. Some participants felt that their emotional struggles impacted their older children.

### Important Aspects of Care Across Antepartum and Postpartum Periods

#### Peer Support from Others with Similar Experiences

Participants expressed a desire for opportunities to discuss their experiences with other people who have been through something similar. Some commented that they would not have been interested in peer support from women with uncomplicated pregnancies, only women with comparable diagnoses or a similar level of acuity.

## Discussion

In this qualitative study, former patients who delivered with PAS shared their emotional experiences and offered feedback on health care services that were helpful or would have possibly improved their experience. Findings can be used to guide the integration of additional support into high-risk obstetric practices caring for patients with potentially morbid delivery experiences. Several important takeaways can be drawn from our findings.

### Patients’ Emotional Experiences

Regarding patients’ emotional experiences, anxiety-related emotions (fear, anxiety, guilt and trauma) were more common than depression. Specifically, fear and isolation emerged in the antepartum period, while grief, anxiety and trauma were emphasized in the postpartum period. This is consistent with other PAS-focused studies that showed elevated rates of fear^18^, trauma^9,17,18^ and isolation^17^. Notably, postpartum depression (PPD) was not a prominent theme in the current study. Parallel findings were shown by Grover et al, who found that patients with PAS had more anxiety than those without PAS six months following delivery, but not more depression. However, both symptoms were elevated at later time points.^23^ Similarly, a past study on mental health experiences of patients with non-PAS pregnancy complications (preeclampsia or premature membrane rupture) showed higher rates of postpartum PTSD, but not depression, when compared to uncomplicated obstetric patients.^3^

While most obstetric practices are equipped to screen and initiate care for PPD, the complex symptoms of fear, grief, and PTSD are outside of most obstetricians’ expertise. Integrating members of the care team who can recognize and manage these symptoms is therefore crucial to comprehensive high-risk obstetric care involving morbid deliveries. Additionally, enhancing patient education about the prevalence of postpartum anxiety- and stressor-related disorders rather than focusing primarily on PPD, may be helpful for building awareness among patients and their families.

### Data-Driven Health Services Recommendations

Our results support several recommendations for providing effective integrated mental health services for high-risk obstetric conditions. First, mental health support should be presented as standard care, following an “opt-out” rather than “opt-in” approach. Patients may have declined mental health services during or after their pregnancies, but in hindsight recognized that it would have been beneficial.

Second, the care team should consist of providers well informed on the high-risk obstetric condition – in the current case, “accreta-informed” providers. Consistent care from knowledgeable providers can build trust and confidence in the care team and avoid burdening patients with educating their providers. Involving mental health specialists in multidisciplinary patient care meetings will help providers to understand what individual patients are facing, with the added benefit of informing the full care team of a patient’s emotional needs.

Third, practicing trauma-informed care (TIC) may be essential to mitigate high-risk patients’ experiences. This includes recognizing patients’ emotional states, offering consistent messaging and care provision, giving patients a sense of agency in their care decisions, and appropriately conducting discussions of delivery events. Our patients requested longer postpartum follow-up, acknowledging that early discussions with their providers may have been ineffective. This aligns with the literature around TIC suggesting that survivors may need temporal distance from a traumatic event before being able to process it.^24^ Mental health specialists can play an important role in educating the entire care team on TIC and promoting these practices on a patient-by-patient basis.

Fourth, consistent with two of the recommendations from a prior study^16^, our finding support offering family-centered care and providing consistent providers for their longer-term follow-up. This means offering services not only to the obstetric patient, but also to the partner and/or other key family members and potentially offering family therapy sessions. Our findings suggest that these services would have a direct impact on the short- and long-term health of the obstetric patient. Longer follow-up times means providing wrap-around services for at least a year or longer for patients with complicated pregnancies and deliveries. As above, evidence-based treatments for trauma are typically not recommended until initial stabilization in the immediate aftermath of a traumatic experience. It is possible that in the context of the postpartum period, this initial stabilization period is longer than it would be in other contexts.

Fifth, our findings suggest that beyond giving information about mental health support^16^, fully incorporating mental health expertise into the multidisciplinary obstetrics care team is the optimal means to bring needed services to these patients. Many practices will lack the available resources to integrate a regular team of mental health professionals into their obstetric care teams, but this goal should be prioritized in supporting patients with complicated pregnancy or postpartum conditions or morbid deliveries. In the setting of resource limitations, obstetric providers can still support their patients by educating community mental health providers to whom they refer their patients; consulting with outside providers when they become involved in their patients’ care; and bringing condition-specific, TIC education to their care teams.

### Limitations

One limitation of the study was selection bias. Patients who were not comfortable sharing their experiences in a group format are not captured well by this study. Similarly, the focus group format may select individuals who prefer group and peer support, which would not apply to all patients with comparable experiences. Additionally, though the recruitment process was blinded to race and ethnicity, the ultimate study population’s demographic breakdown had no representation from Black or Asian patients, who represent respectively 13% and 10% of the hospital’s confirmed PAS patients. It is important that future research in PAS and traumatic birth continues to proactively seek out viewpoints from underrepresented populations, including by considering alternate approaches to study design (such as individual interviews or surveys). Finally, retrospective design and recall bias may also have played a role, as many participants were several years out from their delivery.

## Conclusions

Using placenta accreta spectrum as a specific example of high-morbidity deliveries, we observed that patients anticipating and undergoing these deliveries experience prominent anxiety and trauma-related negative emotions. Introducing well-informed, well-integrated mental health experts into multidisciplinary care teams should improve recognition, management, and ideally mitigate these adverse emotional experiences. An optimal model should include routine, trauma-informed, family-centered, long-term follow-up care for these patients.

## Data Availability

All data produced in the present study are available upon reasonable request to the authors.

## List of Abbreviations

PTSD: Post-traumatic stress disorder
PAS: Placenta accreta spectrum
BWH: Brigham and Women’s Hospital
PPD: Postpartum depression
TIC: Trauma-informed care

## Declarations

### Ethics approval and consent to participate

Approval was received from the Mass General Brigham Institutional Review Board prior to conducting the study (IRB#2018P000307). Eligible participants received written information about the study, including the use of audio recording and deidentification of transcripts. Informed consent to participate was obtained from all participants in the study prior to participation in the focus groups. This study adhered to the Declaration of Helsinki.

### Consent for publication

Not applicable

### Availability of data and materials

The interviews transcribed for the present study are not publicly available due to privacy considerations. Following possible approval by the Mass General Brigham Institutional Review Board, the corresponding author could make deidentified transcripts available upon reasonable request.

### Competing interests

The authors declare that they have no competing interests.

### Funding

Financial Support was received through an unrestricted donation from the Hess Foundation, which was not involved in study design or recruitment; data collection, analysis, or interpretation; manuscript preparation; or decision to publish.

### Authors’ contributions

NF coded and interpreted the data and took a lead role in manuscript preparation; MN coded and interpreted the data and took a lead role in manuscript preparation; JL contributed to the design of the analysis plan and participated in data interpretation and manuscript preparation; KS coded and interpreted the data and participated in manuscript preparation; LC developed the focus group questions, conducted the focus groups, and participated in data interpretation; PW conducted all patient recruitment and communications, oversaw transcript production and deidentification, and maintained study databases; LM served as the principal investigator, designed the study, developed focus group questions, conducted the focus groups, and participated in data interpretation; DC served as the co-investigator, designed the study, developed focus group questions, participated in final data interpretation, and provided major edits to the manuscript. All authors read and approved the final manuscript.

## Acknowledgements

Not applicable.

## Notes

### Competing Interest Statement

The authors have declared no competing interest.

### Author Declarations

Ethics committee/IRB of Mass General Brigham gave ethical approval for this work.

