## Supplementary File 1 for "Integrating mental health support into care for placenta accreta spectrum: A qualitative analysis of patient perspectives"

**Supplementary File 1: Questions for Traumatic Birth Focus Group**

1. What were the aspects of care DURING pregnancy that were helpful?
    a. Were there staff or particular people who were most helpful during prenatal care

2. Are there things that could have been added to your care that would have been helpful to your emotional wellbeing?
    a. Were there particular times during prenatal care when you felt there was more help needed?
    b. Were there times during your hospital stay(s) that you felt more could have been done?
    c. Were there times postpartum?
    d. Would longer standing connection postpartum be helpful?
    e. What resources would you like to see added to enhance emotional wellbeing during pregnancy and postpartum?

3. What are the impacts that your complicated pregnancy and delivery had on your mental and emotional wellbeing?
    a. When did you notice this impact?
    b. How long did it persist?
    c. What (if any) mental health care did you pursue?
    d. Did you find peer support? Would that be important to include?
    e. What aspects of your life did this affect?
    f. Would mental health resources at BWH be helpful?

4. Would you have been interested in mental health care as part of your obstetric care – either during or after birth?
    a. What type of MH resources – group therapy, indiv therapy, medications?
    b. Where would you like to see such resources offered? Would it be helpful to have them in the same location as OB care or would it be better to be in a separate place?

5. Did you find resources outside your care at BWH that were particularly helpful (such as other providers, groups, internet resources, peers/family)?
    a. Could any of these things be incorporated into the care here?

6. In addition to the medical issues, were there other factors that were particularly stressful during your pregnancy, or contributed to the stress of your experience?
    a. During what phase of care (prenatal, postpartum)?
    b. What can be done to address these stressors?

7. How was your family impacted by the experience you had during your pregnancy and delivery?
    a. What resources would have helped them?
